# A novel algorithm to detect non-wear time from raw accelerometer data using convolutional neural networks

**DOI:** 10.1101/2020.07.08.20148015

**Authors:** Shaheen Syed, Bente Morseth, Laila A. Hopstock, Alexander Horsch

## Abstract

Current non-wear detection algorithms frequently employ a 30- to 90-minute interval in which recorded acceleration needs to be below a threshold value. Such intervals need to be long enough to prevent false positives (type I errors), while short enough to prevent false negatives (type II errors), limiting their ability to achieve a high F1 score. In this paper, we propose a novel non-wear detection algorithm that eliminates the need for an interval. Rather than inspecting acceleration within intervals, we explore acceleration patterns right before and right after an episode of non-wear time. By drawing on insights from the field of activity type recognition, we propose an algorithm that uses a convolutional neural network to detect the preceding activity ‘taking off the accelerometer’ and the following activity ‘placing it back on’. We evaluate our algorithm against several baseline and existing non-wear algorithms for raw accelerometer data, and our algorithm achieves a perfect precision, a recall of 0.9962, and an F1 score of 0.9981, outperforming all evaluated baseline and non-wear algorithms. Although our algorithm was developed using patterns learned from a hip-worn accelerometer, we propose algorithmic steps that can easily be applied to a wrist-worn accelerometer and a retrained classification model.

## 1 Introduction

Accelerometer-based motion sensors have become a popular tool to measure and characterise daily physical activity (PA)^1–4^. The use of accelerometers in research and consumer applications has grown exponentially^5^, as accelerometers offer versatility, minimal participation burden, and relative cost efficiency^6–8^. As a result, accelerometers have become the standard tool for measuring PA in large epidemiological cohort studies^9^.

One essential step in the processing of accelerometer data is the detection of the time the accelerometer is not worn (non-wear time)^10^. Non-wear time can occur during sleep, sport, showering, water-based activities, or simply when forgetting to wear the accelerometer. Non-wear detection algorithms developed for count-based accelerometer data typically look for periods of zero acceleration within specified time intervals, such as 30, 60, or 90 minutes^11–13^. Unfortunately, the accuracy of current count-based non-wear algorithms is sub-optimal as they frequently misclassify true wear time as non-wear time (type I error)^14^, especially during episodes of sleep and sedentary behaviour^15–18^.

During recent years, with technological advances, accelerometers are able to record and store raw acceleration data (in gravity units [*g*]) over three axes with sample frequencies up to 100Hz or more^5^. The use of raw data opens up new analytical methods and, in contrast to count-based methods^8^, could enable a direct comparison of the data obtained from different accelerometer devices^5^. However, the development of non-wear algorithms for raw acceleration data has received little attention, despite the widespread adoption of raw accelerometer sensors in PA related studies. These algorithms typically examine the standard deviation (SD) and value ranges of the acceleration axes within a certain time interval and associate low values with non-wear time^19,20^. In addition, a recent study has evaluated and proposed other means of determining non-wear time, such as inspecting acceleration values when filtering the data (high-pass filter), or by inspecting changes in tilt angles (slope)^21^.

However, all current algorithms employ a rather long minimum time interval (e.g. 30 minutes or 60 minutes) in which a specific measure (e.g. the SD, vector magnitude unit (VMU) or tilt) needs to be below a threshold value. The underlying rationale for using such a time interval is arguably based on analytic approaches adopted from traditional count-based algorithms^5^. A major drawback of algorithms employing a time interval is that any non-wear episode shorter than the interval cannot be detected. This would negatively impact the recall (also referred to as sensitivity) performance and can cause an increase in false negatives or type II errors; true non-wear time inferred as wear time. In other words, it is rather safe to assume that an interval of 60 minutes of no activity can be considered non-wear time, albeit that this assumption comes at a cost.

To remedy the above, and to fully unlock the potential of raw accelerometer data, this paper explores an analytical method frequently employed in activity type recognition studies. That is, the use of deep neural networks to detect activity types such as jogging, walking, cycling, sitting, and standing^22–27^, as well as more complex activities such as smoking, eating, and falling^28,29^. Following this line of research, we hypothesise that episodes of non-wear time precede and follow specific activities or movements that can be characterised as taking off the accelerometer and placing it back on, and that such activities can be detected through the use of deep neural networks. In doing so, we can distinguish episodes of true non-wear time from episodes that only show characteristics of non-wear time, but are in fact wear time.

We utilised a gold-standard dataset with known episodes of wear and non-wear time constructed from two accelerometers and electrocardiogram (ECG) recordings^14^. We trained several convolutional neural networks to classify activities that precede and follow wear and non-wear time. In doing so, we aimed to develop a novel algorithm to detect non-wear time from raw acceleration data that can detect non-wear time episodes of any duration, thus removing the need for currently employed time intervals. To evaluate the performance of our algorithm, we compared it with several baseline and previously developed non-wear algorithms that work on raw data^19–21^.

## 2 Methods

### 2.1 Gold-standard dataset

The gold-standard dataset was constructed from a dataset containing raw accelerometer data from 583 participants of the Tromsø Study, a population-based cohort study in the municipality of Tromsø in Norway, and includes seven data collection waves taking place between 1974 and 2016^30,31^. Our dataset was acquired in the seventh wave of the Tromsø Study. Tromsø 7 was approved by the Regional Committee for Medical Research Ethics (REC North ref. 2014/940) and the Norwegian Data Protection Authority, and all participants gave written informed consent. The usage of data in this study has been approved by the Data Publication Committee of the Tromsø Study. Furthermore, all methods were carried out in accordance with relevant guidelines and regulations (i.e. Declaration of Helsinki).

The dataset contains, for each of the 583 participants, raw acceleration data recorded by an ActiGraph model wGT3X-BT accelerometer (ActiGraph, Pensacola, FL) with a dynamic range of *±*8 *g* (1*g* = 9.81 m·s^−2^). The ActiGraph recorded acceleration in gravity units *g* along three axes (vertical, mediolateral and anteroposterior) with a sampling frequency of 100Hz. In addition, this dataset contains data from the simultaneously worn Actiwave Cardio accelerometer (CamNtech Ltd, Cambridge, UK) with a dynamic range of *±*8 *g* that recorded raw acceleration data along three axes, as well as a full single-channel ECG waveform. The dataset consisted of data from 267 (45.8%) males and 316 (54.2%) females aged 40–84 (mean = 62.74; SD = 10.25). The participants had a mean height of 169.81 cm (SD = 9.35), a mean weight of 78.31 kg (SD = 15.27) and a mean body mass index of 27.06 kg/m^2^ (SD = 4.25).

Based on this dataset, a gold-standard dataset with labelled episodes of true non-wear time was constructed by training a machine learning classifier that focused on discrepancies between the various signals. The procedure is explained in detail in our previous study^14^, and also details information regarding the frequency of non-wear episodes, their duration and distribution over the course of a day. The constructed gold-standard dataset contains start and stop timestamps for episodes of true non-wear time derived from raw triaxial 100Hz ActiGraph acceleration data, and will serve as ground truth labels in subsequent steps of our proposed algorithm. Henceforth, the Actiwave Cardio data has not been used since it was only used in the construction of the gold-standard dataset.

### 2.2 Finding candidate non-wear episodes

The proposed raw non-wear detection algorithm works on the basis of candidate non-wear episodes that are defined as episodes of no activity that show characteristics of true non-wear time but cannot yet be classified as true non-wear time. Candidate episodes occur during actual non-wear time, in which a candidate episode becomes an episode of true non-wear time, but they can also occur during sedentary behaviour or sleeping since the accelerometer records no movement for a certain amount of time. For illustrative purposes, acceleration data with several candidate non-wear episodes are shown in the Supplementary Fig. S1.

Candidate non-wear episodes were detected by calculating the SD of the raw triaxial data for each 1-minute interval. By visual inspection of the data, a SD threshold of ≤ 4.0 m*g* (0.004 *g*), recognisable by horizontal or flat plot lines, was found appropriate to obtain candidate non-wear episodes. More concretely, lowering this threshold would not detect any episode of physical inactivity, meaning that 4.0 m*g* is very close to the accelerometer’s noise level. Consecutive 1-minute intervals were grouped into candidate non-wear episodes. Additionally, a forward and backward pass over the acceleration data for each of the candidate non-wear episode were performed to detect the edges on a 1-second resolution, that is, the exact point in which an episode of no activity (i.e. SD > 4.0 m*g*) follows or precedes some activity (i.e. SD ≤ 4.0 m*g*). For each candidate episode, the exact start and stop timestamp on a 1-second resolution was recorded.

### 2.3 Creating features

The next step in the construction of the non-wear algorithm was to detect the activity associated with *taking off the accelerometer* and *putting the accelerometer back on*, from background activity (i.e. activity that occurs before and after a candidate non-wear episode that is not true non-wear time). In doing so, we extracted a segment or window of raw triaxial acceleration data right before (i.e. preceding) and right after (i.e. following) a candidate non-wear episode, and used the raw triaxial acceleration data as features.

Different features were created by varying the window size from 2–10 seconds, since the optimal window size was unknown at that point. Technically, a preceding feature is extracted from *t*_*start*_ − *w* up to *t*_*start*_, where *t*_*start*_ is the start timestamp of the episode in seconds and *w* is the window size in seconds. The following feature is extracted from *t*_*stop*_ until *t*_*stop*_ + *w*, with *t*_*stop*_ marking the end of an episode; for example, a preceding feature with a 5-second window would yield a (100Hz × 5s) by (3 axes) = (500 × 3) matrix. In addition, by utilising our labelled gold-standard dataset, each constructed feature was either given the label 0, if it preceded or followed wear time, or 1, if it preceded or followed non-wear time; no differences were made between start and stop events. To illustrate this, Fig. 1 displays several start and stop segments from candidate non-wear episodes where the features were extracted. Importantly, no additional filtering or pre-processing was performed on the raw data, and features belonging to the minority classes were up-sampled by random duplication so as to create a class balanced dataset.

**Figure 1.**
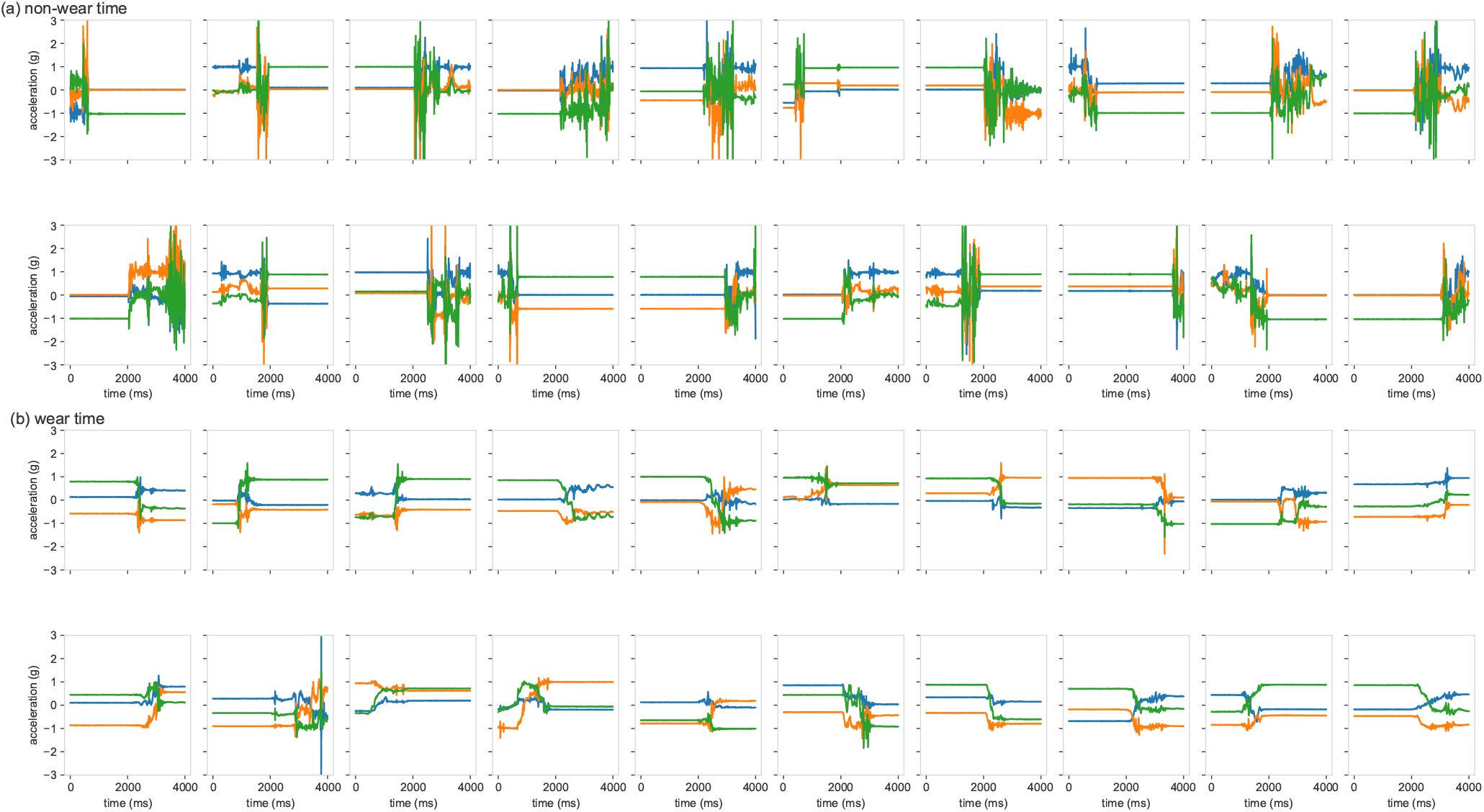
Start or the stop segments of candidate non-wear episodes where features of a length of 2–10 seconds were extracted; (**a**) start or stop episodes of true non-wear time, (**b**) start or stop episodes of wear time.

### 2.4 Training a deep neural network

Convolutional neural networks (CNN) are designed to process data in the form of multiple arrays^32^. CNNs are able to extract the local dependency (i.e. nearby signals that are likely to be correlated) and scale invariant (i.e. scale-invariant for different paces or frequencies) characteristics from the feature data^27^. The 1-dimensional (1D) CNN is particularly suitable for signal or sequence data such as accelerometer data^32^ and, to date, 1D CNNs have successfully been applied for human activity recognition^28,33^, and outperform classical machine learning models on a number of benchmark datasets with increased discriminative power^34^.

A total of four 1D CNN architectures were constructed and trained for the binary classification of our features as either belonging to true non-wear time or to wear time episodes. Figure 2 shows the four proposed architectures labelled V1, V2, V3, and V4. The input feature is a vector of *w* × 3 (i.e. three orthogonal axes), where *w* is the window size ranging from 2–10 seconds (note that a single second contains 100 datapoints). In total, 10 × 4 = 40 different CNN models were trained. CNN V1 can be considered a basic CNN with only a single convolutional layer followed by a single fully connected layer. CNN V2 and V3 contain additional convolutional layers with different kernel sizes and numbers of filters. Stacking convolutional layers enables the detection of high-level features, unlike single convolutional layers. CNN V4 contains a max pooling layer after each convolutional layer to merge semantically similar features while reducing the data dimensionality^32^. A CNN architecture with max pooling layers has shown varying results, from increased classification performance^33^ to pooling layers interfering with the convolutional layer’s ability to learn to down sample the raw sensor data^34^. All proposed CNN architectures have a single neuron in the output layer with a sigmoid activation function for binary classification.

**Figure 2.**
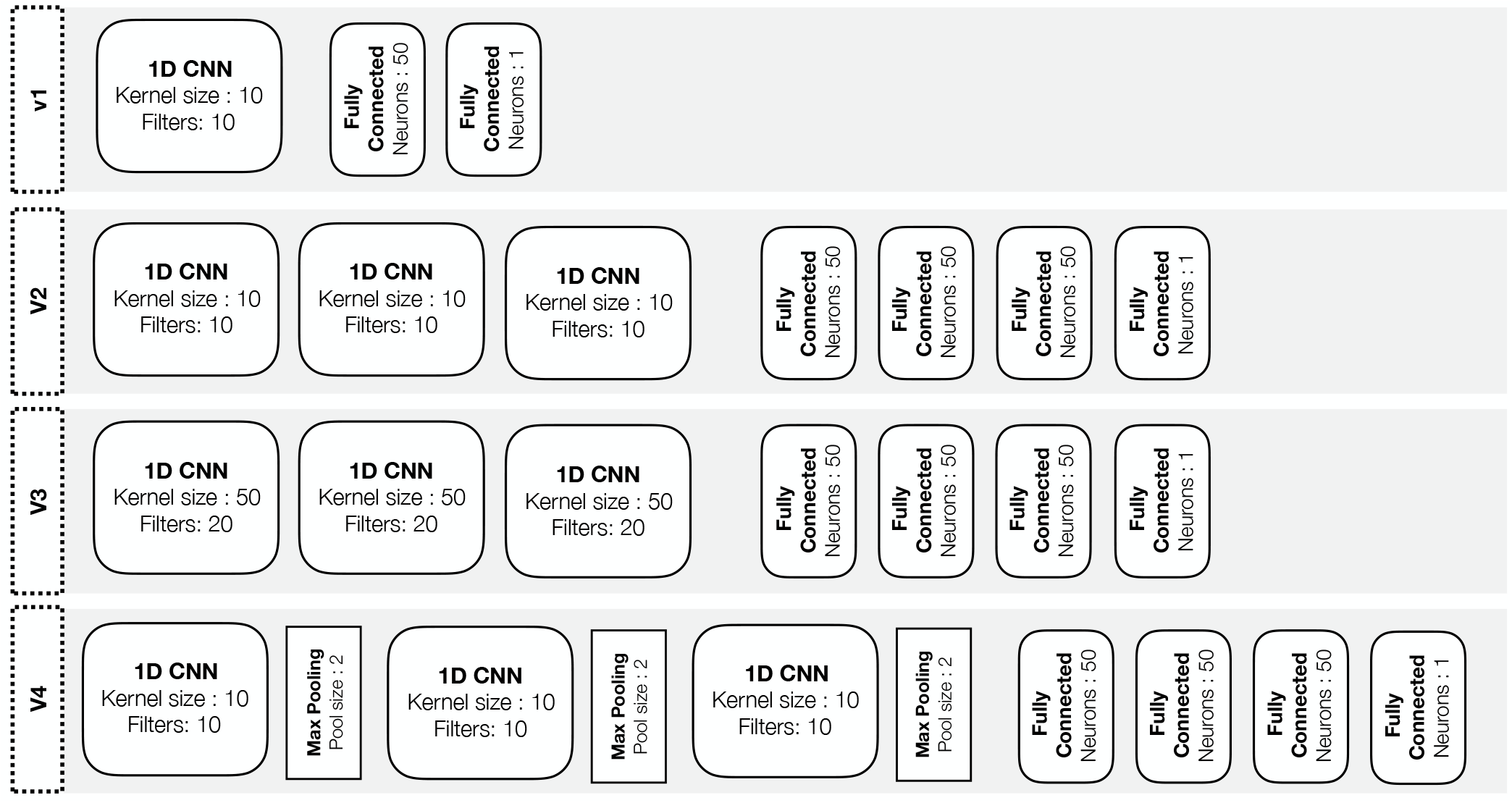
Overview of the four convolutional neural network architectures used for binary classification of start and stop features extracted from wear and non-wear episodes. Before the first fully connected layer, the output data from the previous layer is flattened.

Training was performed on 60% of the data, with 20% used for validation and another 20% used for testing. All models were trained for up to 250 epochs with the Adam optimiser^35^ and a learning rate of 0.001. Loss was calculated with binary cross entropy and, additionally, early stopping was implemented to monitor the validation loss with a patience of 25 epochs and restore weights of the lowest validation loss. This means that training would terminate if the validation loss did not improve for 25 epochs, and the best model weights would be restored. All models were trained on 2 × Nvidia RTX 2080TI graphics cards and programmed in the Python library TensorFlow (v2.0)^36^.

### 2.5 Inferring non-wear time from raw acceleration data

At this stage, the trained CNN model can only classify the start and stop windows of a candidate non-wear episode. To fully detect non-wear episodes from raw acceleration data, the following four steps were applied to the algorithm.

#### Detecting candidate non-wear episodes

As discussed in the previous Section, detecting candidate non-wear episodes was based on a forward pass through the raw acceleration data to detect 1-minute intervals in which the acceleration has a SD of ≤ 0.004 *g*. Consecutive 1-minute intervals below this threshold are merged together and considered a single candidate non-wear episode; these episodes formed the basis of the non-wear detection algorithm.

#### Merging bordering candidate non-wear episodes

Due to artificial movement, a potentially longer non-wear episode might be broken up into several candidate non-wear episodes that are in close proximity to each other. More concretely, the forward search for 1-minute intervals with ≤ 0.004 *g* SD threshold would not include the 1-minute interval in which artificial movement (i.e. a spike in the acceleration) occurred. As a result, when merging together consecutive 1-minute intervals, the artificial movement stops this consecutive sequence. The duration of the artificial movement can also vary; for example, moving the sensor from the bathroom to the bedroom can take multiple seconds, whereas a nudge or touch while the accelerometer lies on a table or nightstand can be a single second. The first hyperparameter of the algorithm defines the merging length in seconds, and five different values of 1, 2, 3, 4, and 5 seconds were explored; for example, a merging length of 3 seconds means that two candidate non-wear episodes that are no more than 3 seconds apart are merged together into a single longer candidate non-wear episode.

#### Detecting the edges of candidate non-wear episodes

Candidate non-wear episodes were detected with a minute resolution (i.e. by using a 1-minute interval). However, as it was necessary to determine what happened immediately before (preceding) or immediately after (following) an episode, this resolution was too low. To find the exact timestamp when a candidate non-wear episode started and stopped, a forward and backwards search on a resolution of 1-second was performed. More concretely, the edges were incrementally extended, and the SD was calculated for each extended 1-second interval. When it remained ≤ 0.004 *g*, the search was continued. This was done forwards to find the exact end of an episode, and backwards to find the exact start of an episode.

It is important to note that the detection of candidate non-wear episodes could have been performed with 1-second intervals, rather than with 1-minute intervals. The latter, however, is computationally faster and eliminated the detection of a high number of unwanted candidate non-wear episodes that were only a few seconds in duration.

#### Classifying the start and stop windows

Activity preceding the start of a candidate non-wear episode was extracted with a window length of 2–10 seconds, as well as activity that followed from the end of the candidate non-wear episode with a window length of 2–10 seconds. The exact window length was dependent on the best F1 classification performance measured on the (unseen) test set of the CNN model constructed and described in the previous Section. After class inference of both the start and stop activity, two logical operators `AND` and `OR` were inspected to determine if both sides or a single side resulted in a better detection of true non-wear time. This logical operator was the second hyperparameter to be optimised, and it can take on two different values: `AND` for both sides and `OR` for a single side. In addition, candidate non-wear episodes can occur at the start or the end of the acceleration signal and, as such, a preceding or following window cannot be extracted since there is no data. It was also investigated if such cases should default to wear or non-wear time. Default classification for the beginning or end of the activity data was the third hyperparameter to be optimised and this could default to two different values: non-wear time or wear time.

A total of 5 × 2 × 2 = 20 different combinations of hyperparameter values were tested to explore their classification performance on the gold-standard dataset. To prevent these parameterisations from overfitting to the dataset, their performance on a random sample of 50% (training set) of the participants from our gold-standard dataset was explored, that is *n* = 291, as well as their classification performance on the remaining unseen 50% (test set) of the participants. In doing so, the aim was to provide hyperparameter values that can generalise to other datasets.

### 2.6 Calculating classification performance

The classification performance is calculated when applying the CNN classification model and the steps described in the previous Section to the gold-standard dataset comprising raw acceleration data from 583 participants. The following classification metrics were reported on: accuracy, precision, specificity, recall (analogous to sensitivity), and F1. The false positives, true positives, false negatives, and true negatives were calculated by looking at 1-second intervals of the acceleration data and comparing the inferred classification with the gold-standard labels. This process is graphically displayed in Supplementary Fig. S2.

### 2.7 Evaluating classification performance

Our proposed non-wear algorithm was evaluated against the performance of several baseline and existing non-wear detection algorithms^19,20^.These baseline algorithms employ a similar analytical approach commonly found in count-based algorithms^11–13^, that is, detecting episodes of no activity by using an interval of varying length.

The first baseline algorithm detected episodes of no activity when the acceleration data of all three axes had a SD threshold of ≤ 0.004 *g*, ≤ 0.005 *g*, ≤ 0.006 *g*, and ≤0.007 *g* and the duration did not exceed an interval length of 15, 30, 45, 60, 75, 90, 105, or 120 minutes. A similar approach was proposed in another recent study as the *SD_XYZ* method^21^, although the authors fixed the threshold to 13 m*g* and the interval to 30 minutes for a wrist worn accelerometer. Throughout this paper, the first baseline algorithm is referred to as the *XYZ* algorithm.

The second baseline algorithm was similar to the first baseline algorithm, albeit that the SD threshold was applied to the vector magnitude unit (VMU) of the three axes, where VMU is calculated as 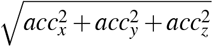, with *acc*_*x*_, *acc*_*y*_, and *acc*_*z*_ referring to each of the orthogonal axes. A similar approach has recently been proposed as the *SD_VMU* algorithm^21^. Throughout this paper, this baseline algorithm is referred to as the VMU algorithm.

Last, our algorithm is evaluated against the Hees algorithms (details of which can be found in the open source library GGIR^37^) with a 30 minute interval^19^, a 60 minute interval^20^, and a version with optimised hyperparameters and a 135 minute interval. Throughout this paper, these three algorithms are referred to as *HEES_30, HEES_60*, and *HEES_135*, indicating their interval length in minutes. Additionally, the sliding window used in the Hees algorithms has been lowered to 1 minute, instead of the default 15 minutes, to make it similar to the sliding window used in the other evaluated algorithms.

## 3 Results

### 3.1 Convolutional neural network

Figure 3 presents the classification performance of the four evaluated CNN architectures. The V2 CNN architecture obtains near perfect F1 scores on the training (60%), validation (20%) and test set (20%) with window sizes ranging from 3–7 seconds. Also, with similar training, validation, and test scores, the models show no signs of overfitting to the training data. The V2 architecture also outperforms the V1, V3, and V4 proposed architectures in terms of accuracy, precision, recall, and F1. The simpler V1 architecture—consisting of a single convolutional layer and a single fully connected layer—outperforms the more complex architectures V3 and V4, though all architectures were trained for a sufficient number of epochs. This improvement holds for all datasets (training, the validation, and test set). The V4 architecture, that implements max pooling layers as a way to downsample the features and is otherwise identical to V2, does not seem to perform as well as the V2 architecture in terms of performance measured during the training, validation, and test sets.

**Figure 3.**
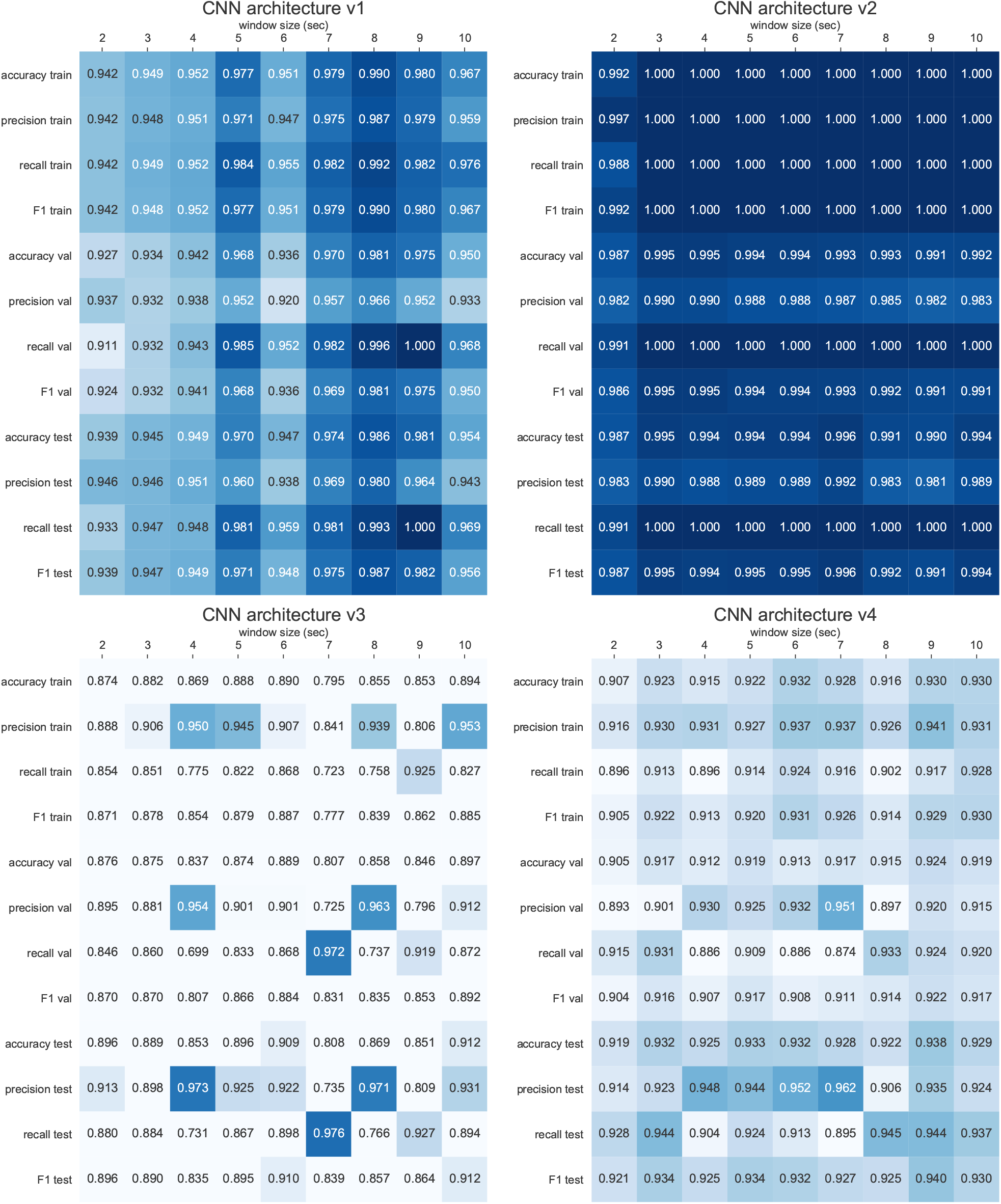
Accuracy, precision, recall, and F1 performance metrics for training data (60%), validation data (20%), and test data (20%) for the four architectures evaluated. All CNN models were trained for a total of 250 epochs with early stopping enabled, a patience of 25 epochs, and restoring of the best weights when the validation loss was the lowest.

Looking at the V2 architecture, a window size of 3 seconds provides a marginal increase in F1 performance on the test set (0.995) when compared to the 2 second window (0.987). Further increasing the window to 7 seconds shows a very minor increase in F1 performance on the test set (0.996), however, taking into account the 95% confidence interval for the 7 second window (*±* 0.00512), the difference between the CNN models with a 3–6 second window is not statistically significant. For the remainder of the results, the V2 CNN model with a window size of 3 seconds was selected as the CNN model to infer if start and stop segments of candidate non-wear time belong to true non-wear time or to wear time episodes. Furthermore, by using a 3-second window, compared to a 7-second window, there is a reduction in the input feature dimensions from (700 × 3 axes) to (300 × 3 axes) for 100Hz data, resulting in the CNN model having 144,031 parameters instead of 344,031. An overview of the training and validation loss, including the performance metrics accuracy, precision, recall, F1, and area under the curve (AUC) are presented in the Supplementary Fig. S3.

### 3.2 Non-wear time algorithm hyperparameters

The ability to classify start and stop segments of a candidate non-wear episode is one function of the proposed algorithm. As outlined in Methods Section, there are remaining steps that involve traversing the raw triaxial data and handling the following cases: (i) artificial movement by merging neighbouring candidate non-wear episodes; (ii) inspecting two logical operators `AND` and `OR` to determine if the start and stop segments combined (i.e. `AND`) or a single side (i.e. `OR`) results in a better detection of true non-wear time; and (iii) candidate non-wear episodes at the beginning or end of the acceleration signal that have no preceding start or following end segment. Table 1 presents the classification performance of detecting true non-wear time episodes from a random sample of 50% (i.e. training data) of the participants from the gold-standard dataset when utilising the CNN v2 architecture with a window size of 3 seconds and exploring 20 combinations of hyperparameter values.

**Table 1.** The classification of accuracy, precision, recall, and F1 performance metrics when applying the new algorithm on 50% of the available data (*n* = 291/583) while exploring 20 combinations of hyperparameter values; 95% confidence intervals are shown between parentheses. Merge (mins) = the merging of neighbouring candidate non-wear episodes to handle artificial movement. Logical operator = `AND` if both start and stop segments or `OR` if only one side of a candidate non-wear episode needs to be classified as true non-wear time to subsequently classify the candidate non-wear episode as an episode of true non-wear time. Edge default = the default classification of a candidate non-wear episode that has no start or end segment, such cases that occur right at the beginning or end of the acceleration data and default to wear or non-wear time.

| Merge (mins) | Logical Operator | Edge default | Accuracy | Precision | Recall | F1 |
| --- | --- | --- | --- | --- | --- | --- |
| 5 | AND | non-wear time | 1.0 ($\pm 0.0003$) | 0.9995 ($\pm 0.0019$) | 1.0 ($\pm 0.0$) | 0.9997 ($\pm 0.0013$) |
| 4 | AND | non-wear time | 0.9997 ($\pm 0.0013$) | 0.9995 ($\pm 0.0019$) | 0.9889 ($\pm 0.0085$) | 0.9941 ($\pm 0.0062$) |
| 4 | OR | wear time | 0.999 ($\pm 0.0026$) | 0.9551 ($\pm 0.0168$) | 0.9993 ($\pm 0.0022$) | 0.9767 ($\pm 0.0123$) |
| 3 | OR | wear time | 0.9989 ($\pm 0.0026$) | 0.9552 ($\pm 0.0168$) | 0.9979 ($\pm 0.0038$) | 0.9761 ($\pm 0.0124$) |
| 5 | OR | wear time | 0.9989 ($\pm 0.0027$) | 0.9498 ($\pm 0.0177$) | 1.0 ($\pm 0.0$) | 0.9742 ($\pm 0.0129$) |
| 3 | AND | non-wear time | 0.9974 ($\pm 0.0041$) | 1.0 ($\pm 0.0$) | 0.8785 ($\pm 0.0265$) | 0.9353 ($\pm 0.02$) |
| 2 | OR | wear time | 0.9971 ($\pm 0.0044$) | 0.9437 ($\pm 0.0187$) | 0.9203 ($\pm 0.022$) | 0.9319 ($\pm 0.0205$) |
| 1 | OR | wear time | 0.9962 ($\pm 0.005$) | 0.9158 ($\pm 0.0225$) | 0.9037 ($\pm 0.0239$) | 0.9097 ($\pm 0.0233$) |
| 3 | OR | non-wear time | 0.9955 ($\pm 0.0054$) | 0.828 ($\pm 0.0306$) | 0.9979 ($\pm 0.0038$) | 0.905 ($\pm 0.0238$) |
| 2 | OR | non-wear time | 0.9954 ($\pm 0.0055$) | 0.8244 ($\pm 0.0309$) | 0.9971 ($\pm 0.0044$) | 0.9026 ($\pm 0.0241$) |
| 1 | OR | non-wear time | 0.9949 ($\pm 0.0058$) | 0.8184 ($\pm 0.0313$) | 0.9805 ($\pm 0.0112$) | 0.8922 ($\pm 0.0252$) |
| 4 | OR | non-wear time | 0.9948 ($\pm 0.0059$) | 0.8045 ($\pm 0.0322$) | 0.9993 ($\pm 0.0022$) | 0.8913 ($\pm 0.0253$) |
| 5 | OR | non-wear time | 0.9944 ($\pm 0.0061$) | 0.7924 ($\pm 0.0329$) | 1.0 ($\pm 0.0$) | 0.8841 ($\pm 0.026$) |
| 2 | AND | non-wear time | 0.9954 ($\pm 0.0055$) | 1.0 ($\pm 0.0$) | 0.7862 ($\pm 0.0333$) | 0.8803 ($\pm 0.0264$) |
| 1 | AND | non-wear time | 0.994 ($\pm 0.0063$) | 1.0 ($\pm 0.0$) | 0.7202 ($\pm 0.0364$) | 0.8373 ($\pm 0.03$) |
| 5 | AND | wear time | 0.9908 ($\pm 0.0077$) | 0.9991 ($\pm 0.0025$) | 0.5736 ($\pm 0.0401$) | 0.7288 ($\pm 0.0361$) |
| 4 | AND | wear time | 0.9906 ($\pm 0.0078$) | 0.9991 ($\pm 0.0025$) | 0.5625 ($\pm 0.0403$) | 0.7197 ($\pm 0.0365$) |
| 3 | AND | wear time | 0.9888 ($\pm 0.0086$) | 1.0 ($\pm 0.0$) | 0.4763 ($\pm 0.0405$) | 0.6452 ($\pm 0.0388$) |
| 2 | AND | wear time | 0.9887 ($\pm 0.0086$) | 1.0 ($\pm 0.0$) | 0.4742 ($\pm 0.0405$) | 0.6433 ($\pm 0.0389$) |
| 1 | AND | wear time | 0.9873 ($\pm 0.0091$) | 1.0 ($\pm 0.0$) | 0.4082 ($\pm 0.0399$) | 0.5797 ($\pm 0.0401$) |

The best F1 score (0.9997 *±* 0.0013) on the training set was achieved by: (i) merging neighbouring candidate non-wear episodes that are a maximum of 5 minutes apart from each other, (ii) using the logical operator `AND` (meaning both start and stop segments need to be classified as non-wear time to subsequently classify the candidate non-wear episode into true non-wear time), and (iii) default start and stop segments to non-wear time when they occur right at the start or end of the acceleration signal. Using these hyperparameter values on the remaining 50% of our gold-standard dataset (i.e. test data) achieved similar results: accuracy of 0.9999 (*±* 0.0006), precision of 1.0 (*±*0.0), recall of 0.9962 (*±* 0.005), and F1 performance of 0.9981 (*±* 0.0035). In summary, the proposed algorithm is able to achieve near perfect performance on the training and test dataset when detecting non-wear episodes, both in terms of the ability to correctly classify an episode as true non-wear time (high precision), as well as the ability to detect all available non-wear time episodes present in the dataset (high recall).

### 3.3 Non-wear time algorithm steps

Based on the results presented in Table 1, and the steps outlined in Methods Section, the complete algorithm is presented below.

#### Detect candidate non-wear episodes

Perform a forward pass through the raw acceleration signal and calculate the SD for each 1-minute interval of each axis. If the standard deviation is ≤ 0.004 *g* for all axes, record this 1-minute interval as a candidate non-wear interval. After all of the 1-minute intervals have been processed, merge consecutive 1-minute intervals into candidate non-wear episodes and record their start and stop timestamps.

#### Merge bordering candidate non-wear episodes

Merge candidate non-wear episodes that are no more than 5 minutes apart and record their new start and stop timestamps. This step is required to capture artificial movement that would typically break up two or more candidate non-wear episodes in close proximity.

#### Detect the edges of candidate non-wear episodes

Perform a backward pass with a 1-second step size through the acceleration data from the start timestamp of a candidate non-wear episode and calculate the SD for each axis. The same is applied to the stop timestamps with a forward pass and a step size of 1 second. If the SD of all axes is ≤ 0.004 *g*, include the 1-second interval in the candidate non-wear episode and record the new start or stop timestamp. Repeat until the SD of the 1-second interval does not satisfy the SD threshold ≤ 0.004 *g*. This results in the resolution of the edges now being recorded on a 1-second resolution.

#### Classifying the start and stop windows

For each candidate non-wear episode, extract the start and stop segment with a window length of 3 seconds to create input features for the CNN classification model. For example, if a candidate non-wear episode has a start timestamp of *t*_*start*_, a feature matrix is created as (*t*_*start*−*w*_: *t*_*start*_) × 3 axes (where *w* = 3 seconds), resulting in an input feature with dimensions of 300 × 3 for 100Hz data. If both start and stop features (i.e. logical `AND`) are classified (through the CNN model) as non-wear time, the candidate non-wear episode can be considered true non-wear time. If *t*_*start*_ is *t* = 0, or *t*_*end*_ is at the end of the acceleration data, those candidate non-wear episodes do not have a preceding or following window to extract features from, the start or stop can be, by default, classified as non-wear time.

### 3.4 Evaluation against baseline and existing non-wear algorithms

Figure 4 presents the F1 performance of the two baseline algorithms as outlined in Methods Section. Figure S4 and S5 in the Supplementary Information provides, in additional to the F1 scores, the performance metrics accuracy, precision, and recall for the *XYZ* and *VMU* baseline algorithms respectively. As shown in Fig. 4, increasing the SD threshold from ≤ 0.004 *g* to a higher value resulted in an F1 performance loss for both the *XYZ* and *VMU* baseline algorithms. The XYZ algorithm achieved the highest F1 score with a SD threshold of ≤ 0.004 *g* and an interval length of 90 minutes; further increasing the interval to 105 or 120 minutes is associated with lower F1 scores, 0.836 and 0.829 respectively. The use of longer intervals resulted in non-wear episodes that were shorter than the interval to not be detected, which caused the recall score to be lower. At the same time, shortening the interval length resulted in a higher recall but a lower precision score (Supplementary Fig. S4). The optimal F1 score for the *VMU* baseline algorithm (0.839) was achieved with a SD threshold of ≤ 0.004 *g* and an interval length of 105 minutes. The *VMU* algorithm shows a similar pattern to the *XYZ* algorithm with respect to balancing the trade-off between capturing more non-wear time (higher recall) with shorter intervals, at a cost of lowering the precision (Supplementary Fig. S5), or using longer intervals to detect less overall non-wear time (lower recall), in favour of being more certain that the inferred non-wear time is true non-wear time (higher precision).

**Figure 4.**
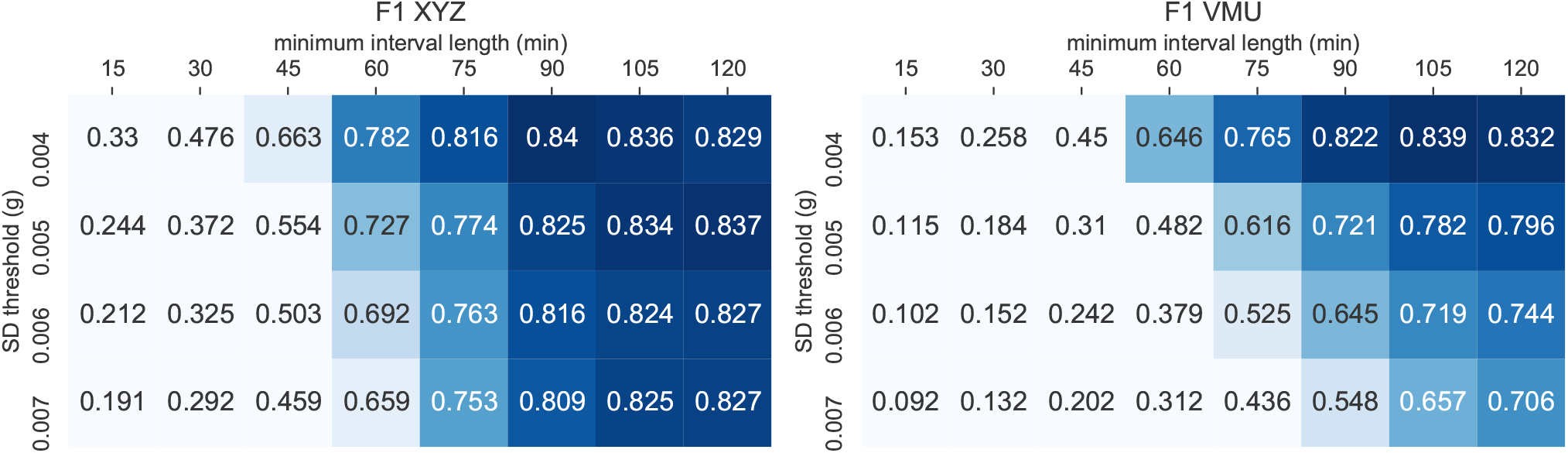
The F1 classification performance of the *XYZ* baseline algorithm (left), and the *VMU* baseline algorithm (right). Note that a SD threshold of 0.003 *g* performed poorly as it is below the accelerometer noise level and is therefore not shown. See Supplementary Fig. S4 and S5 for accuracy, precision, and recall scores.

Besides baseline algorithms, the raw non-wear algorithms developed by van Hees and colleagues with a 30 minute interval^19^ (*HEES_30*), a later published 60-minute interval^20^ (*HEES_60*), and one with a 135-minute interval and tuned hyperparameters^14^ (*HEES_135*) were also evaluated. Figure 5 presents an overview of the obtained classification performance data (precision, recall, and F1) of all evaluated non-wear algorithms against the proposed CNN algorithm. The *HEES_135* algorithm with tuned hyperparameters outperformed the default *HEES_30* and *HEES_60* algorithms with an F1 score of 0.885. In addition, *HEES_135* outperformed the best performing baseline algorithm, *XYZ* (F1 = 0.84) with a 90-minute interval (i.e. XYZ_90), as well as the *VMU* baseline algorithm, (F1 = 0.839) with a 105-minute interval (i.e. *VMU_105*). However, the proposed CNN method outperformed all evaluated non-wear algorithms with a near perfect F1 score of 0.998.

**Figure 5.**
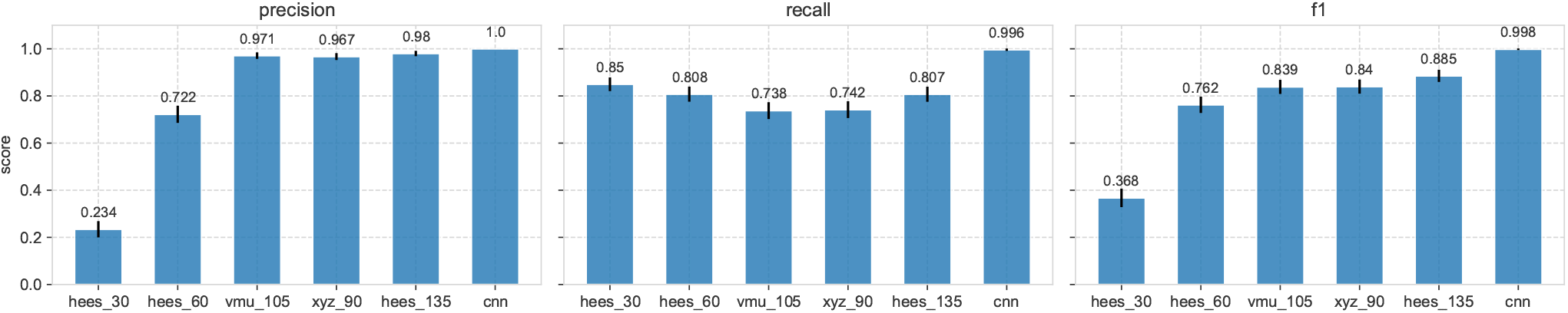
A comparison of the classification performance metrics of the best performing baseline models *XYZ_90* (i.e. calculating the standard deviation of the three individual axes and an interval length of 90 minutes), *VMU_105* (i.e. calculating the standard deviation of the VMU and an interval length of 105 minutes), the *HEES_30* algorithm with a 30 minutes interval, the *HEES_60* with a 60 minutes interval, the *HEES_135* with tuned hyperparameters and a 135 minutes interval, and the proposed CNN algorithm. Error bars represent the 95% confidence interval.

## 4 Discussion

In this paper, we proposed a novel algorithm to detect non-wear time from raw accelerometer data through the use of convolutional neural networks and insights adopted from the field of physical activity type recognition^22–29^. It utilised a previously constructed gold-standard dataset^14^ with known episodes of true non-wear time from 583 participants, and was able to achieve an F1 score of 0.998, outperforming baseline algorithms and existing non-wear algorithms^19,20^.

The main advantage of the proposed algorithm is the absence of a minimum interval (e.g. 30 minutes or 60 minutes) in which a specific metric (e.g. SD) needs to be below a threshold value (e.g. 4 m*g*). Currently, all existing raw and epoch-based non-wear algorithms adopt a minimum interval and have to balance between precision and recall. In other words, a short interval increases the detection of all present non-wear time (higher recall), at the cost of incorrectly inferring wear time as non-wear time (lower precision). Alternatively, a longer interval decreases the detection of all present non-wear time (lower recall), but those detections are more certain to be true non-wear time (higher precision); this trade-off has been discussed at length in our previously published study^14^. A similar finding is shown in Fig. 5, where *HEES_30* achieved a better recall score (0.85) compared to *HEES_60* (0.808) but performed poorly on the precision metric (0.234) in comparison to *HEES_60* (0.772); here both *HEES_30* and *HEES_60* are identical algorithms with the only difference being the interval length.

In line with the above, a larger interval caused a stronger increase in precision scores than a decrease in recall score, subsequently resulting in an overall higher F1 score. In other words, larger intervals perform better in the overall detection and correct classification of both wear and non-wear time; which is what is captured by the F1 metric. In fact, as can be seen from Fig. 5, some of the evaluated baseline and existing non-wear algorithms were able to achieve very high precision scores of 0.971 (*VMU_105*), 0.967 (*XYZ_90*), and 0.98 (*HEES_135*). These results show that the evaluated algorithms can be near perfect in their ability to correctly classify an episode into true non-wear time without too many false positives (type I error). However, a major drawback is that longer intervals cannot detect episodes of non-wear time shorter than the interval. This is a major shortcoming of non-wear algorithms to date and causes their ability to detect all the available non-wear time within the data to be sub-optimal. As a direct consequence, true non-wear episodes shorter than the interval will be inferred as wear time, which can result in an increase of false negatives or type II errors. For datasets with a high frequency of short non-wear time episodes, this can cause derived PA summary statistics to be incorrect, especially summary statistics that are relative to the amount of activity detected. Our proposed CNN algorithm did not perform better on the precision score, however, by not relying on an interval, it was able to detect even the shortest episodes of non-wear time; this enabled the recall score to be high and, as a consequence, resulted in a higher F1 score as well.

### Hyperparameter values

The explored hyperparameter ‘Edge default’ has the risk of being dataset specific, despite our efforts to train on 50% of the data and test on the remaining, unseen, 50%. Its default classification to wear time for episodes without a start or stop segment (those at the beginning or end of the activity data) can be linked to the study protocol and might not translate to other datasets unquestionably. For example, if accelerometers are initialised and start recording before given to the participants, it might be assumed that the recordings after initialisation are non-wear time since the accelerometer still needs to be worn, either shortly thereafter or at a later stage when sent to participants via postal mail. In such cases, a preceding segment can default to non-wear time. However, if accelerometers are initialised to record at midnight (i.e. 00:00), and worn before recording starts, defaulting to non-wear time might not be automatically correct. For example, the participant might sleep before recording starts and, as a result, can obtain a recording at *t* = 0 that does not exceed the ≤ 0.004 *g* SD threshold; in such cases, defaulting to non-wear time would be incorrect.

Additionally, the hyperparameter ‘merge (mins)’, that merges nearby candidate non-wear episodes to capture and include artificial movement, could unintentionally merge true non-wear and wear time episodes if they occur very close to each other (i.e. *<* 5 minutes). For example, if the accelerometer was worn during sleep but removed right after waking up, two candidate non-wear episodes could be detected and merged incorrectly. Although this did not occur in our dataset, reducing the ‘merge (mins)’ hyperparameter to 4 minutes would further reduce the risk of incorrect merging and, given our results, still achieved an F1 score of 0.9941 (*±*0.0062) on our training set (Table 1).

### Limitations and future research

Care must be given to the nature of the PA patterns that we detected in the preceding and following windows of a candidate non-wear episode. As per our results, the CNN model was able to differentiate true non-wear time from wear time segments with near perfect performance based on features taken 3 seconds before the episode started and 3 seconds after the episode ended. Longer feature segments, of 4 or 5 seconds, yielded similar statistical results since, effectively, a shorter segment remains as a subset of a longer segment. The activity patterns of taking off the accelerometer (preceding feature) or putting it back on (following feature) were distinguished from activity patterns that proceeded or followed a candidate non-wear episode that was deemed wear time. In the latter, the CNN model learned patterns that were associated with movement during episodes of no activity, but those where the accelerometer was still worn. Such patterns are, for example, rotating the body during sleep, and during sedentary time, changing sitting positions. All learned patterns were captured from an accelerometer positioned on the right hip. Moreover, the accelerometer was mounted with an elastic waist belt, which could have an additional effect on the learned movement patterns, compared to having a (belt) clip or another way of mounting the accelerometer on the hip. We further suspect the activity patterns to be different for accelerometers positioned on the wrist, since this can be associated with higher movement variability^4^.

A natural next step would thus be to employ a similar approach of detecting non-wear time through activity type recognition by means of deep neural networks for accelerometers positioned at different locations, such as the wrist. With the use of raw accelerometer data, we are confident that our algorithm is invariant to different types of accelerometer brands positioned on the hip, even when data was sampled at different frequencies, as they can easily be resampled to 100Hz^38^. However, other accelerometers may have a different standard deviation threshold value to detect episodes of no activity. Our dataset contained accelerometer data from the ActiGraph wGT3X-BT, which is the most commonly used accelerometer for PA studies^8,17^. A careful analysis revealed that a threshold of 0.004 *g* is close to the accelerometer’s noise level and sufficiently low enough to detect episodes of no activity. This threshold value should, however, be explored for other accelerometers when using our proposed algorithm.

## 5 Conclusion

In this paper, we proposed a novel algorithm that utilises a CNN to detect the activity types of *taking off the accelerometer* and *placing it back on* to enable the detection of non-wear time from raw accelerometer data. Though current raw non-wear time algorithms show promising results in terms of precision scores, their employed interval prevents them from detecting non-wear time shorter than this interval, resulting in a sub-optimal recall score. By classifying activity types, our proposed algorithm does not employ a minimum interval and allows for non-wear time detection of any duration, even as short as a single minute. As per our results, this significantly increased the recall ability and led to a near perfect F1 score (0.998) on our gold-standard test dataset. Although our algorithm was developed for movement associated with a hip-worn accelerometer, future research can be directed at training a CNN for movement associated with a wrist-worn accelerometer, including the optimisation of our algorithm’s hyperparameters.

## Data Availability

The legal restriction on data availability are set by the Tromsø Study Data and Publication Committee in order to control for data sharing, including publication of datasets with the potential of reverse identification of de-identified sensitive participant information. The data can however be made available from the Tromsø Study upon application to the Tromsø Study Data and Publication Committee. Contact information: The Tromso Study, Department of Community Medicine, Faculty of Health Sciences, UiT The Arctic University of Norway.

https://github.com/shaheen-syed/CNN-Non-Wear-Time-Algorithm

https://github.com/shaheen-syed/ActiGraph-ActiWave-Analysis

## Author contributions statement

Study concept and design: S.S. Data collection: B.M., L.A.H. Analysis and interpretation of data: S.S. Drafting of the manuscript: S.S. Critical revision of the manuscript: All authors.

## Additional Information

### Data Availability

The legal restriction on data availability are set by the Tromsø Study Data and Publication Committee in order to control for data sharing, including publication of datasets with the potential of reverse identification of de-identified sensitive participant information. The data can however be made available from the Tromsø Study upon application to the Tromsø Study Data and Publication Committee. Contact information: The Tromsø Study, Department of Community Medicine, Faculty of Health Sciences, UiT The Arctic University of Norway.

All Python code that supports this study is openly available on S.S.’s GitHub page at https://github.com/shaheen-syed/ActiGraph-ActiWave-Analysis

A Python implementation of the CNN non-wear algorithm can be found at https://github.com/shaheen-syed/CNN-Non-Wear-Time-Algorithm

## Supplementary information

Supplementary material accompanies this paper at:

